# Systematic and quantitative analyses of hollow fiber model of *Mycobacterium abscessus* lung disease studies

**DOI:** 10.1101/2025.08.06.25333162

**Authors:** Shashikant Srivastava, Tawanda Gumbo

## Abstract

Guideline-based combination therapy (GBT) achieves sputum culture conversion rates in 23-34% of patients with *Mycobacterium abscessus complex* (MAB) lung disease (LD). Thus, new therapies are needed. We performed a systematic review to validate and benchmark the hollow fiber system model of MAB-LD (HFS-MAB) for drug development. We performed a literature search to identify all published HFS-MAB pharmacokinetics (PK)-pharmacodynamics (PD) studies. Preferred Reporting Items for Systematic Reviews and Meta-Analyses was used for bias minimization. A total of 12 studies were identified. The average quality score was 13.7 out of 21. Eight were monotherapy (exposure-effect and dose-fractionation), one-double β-lactam, and three GBT studies. For omadacycline and imipenem, HFS-MAB data was accompanied by clinical real-world evidence confirming HFS-MAB findings. Monotherapy or combination therapy microbial kill was always terminated by antimicrobial resistance. We used quantitative analyses to rank drugs’ efficacy. The three highest-ranked drugs based cfu/mL fold-kill compared to multi-drug GBT, were sulbactam-durlobactam (177-fold), epetraborole (15-fold), and omadacycline (7-fold). We used the PK/PD target exposures identified by studies in the systematic analysis in Monte Carlo experiments (MCE) to identify optimal doses for inhaled formulations. The optimal inhalational dose of imipenem/cilastatin was 250 mg/day, for tigecycline 4 mg/day, for cefoxitin 50 mg/day, and for amikacin liposome inhalation suspension 590mg once weekly. The HFS-MAB is tractable for exposure-effect, dose-fractionation, and factorial design combination studies. It can be used to rank drugs and inform on which drugs to test in novel combinations. The HFS-MAB fulfills the US Food and Drug Administration Roadmap definition of non-animal New Approach Methodologies.

---

*Mycobacterium abscessus complex* (MAB) is responsible for 12% of all non-tuberculous mycobacteria (NTM) lung disease (LD) in the United States but could contribute up to 42% of NTMs elsewhere (1, 2). In a large study from Spain, MAB *subsp. abscessus* accounted for 52%, MAB *subsp. massiliense* for 34%, and MAB *subsp. bolletii* for 14% of all isolates (3). Guideline-based therapy (GBT) includes an initial phase of treatment of at least three active drugs, based on the susceptibility testing, including one to two from the group of injectables such as amikacin or β-lactams (imipenem or cefoxitin) or tigecycline, and two from orally formulated drugs such as clarithromycin (if no inducible resistance), or clofazimine or linezolid (4). A meta-analysis of 19 clinical studies demonstrated that GBT achieved sputum culture conversion (SCC) in only 34% of patients with MAB *subsp. abscessus* and 54% infected with MAB *subspecies* as initial therapy; for salvage therapy the SCC was 23% for all subspecies (5). This means that it will be important to study these multiple subspecies when developing new therapies. In addition, the US Food and Drug Administration (FDA) released a roadmap of New Approach Methodologies (NAMs) that “offer the tools to assess safety, efficacy, and pharmacology of drugs and therapeutics *without* traditional animal models” (6). The European Medicines Agencies (EMA) has a parallel roadmap (7). We (including Dr. Beatriz Ferro) designed the hollow fiber model (HFS) of MAB-LD (HFS-MAB), which in tandem with Monte Carlo experiments (MCE), fits that designation of NAMs, to test new therapies for MAC-LD (8).

One of the most important factors associated with GBT failure is the presence of cavities, especially those with diameter greater than 2 cm which have odds ratios of 2.4 for cure and 2.5 for death (9, 10). Computed tomography scans demonstrate cavities in 35% of patients at a median diameter of 3 cm (9). Lung cavities and nodules represent a physical barrier to antibiotics, whose concentrations fall across the gradient inversely proportional to the diameter (11, 12). Lung cavities also represent larger MAB burdens, with pre-treatment bacterial burdens (*B_0_*) in the range of 10^7^-10^8^ cfu/lung in patients (10, 13). Large bacterial burdens are associated with poorer outcomes in patients (10, 14–17). Moreover, these MAB burdens are higher than the inverse of the mutation frequencies of GBT drugs, amikacin, clarithromycin, and cefoxitin at 1.8 x 10^-6^ to 4.7 x 10^-7^ cfu/mL (18–21). This means that at the start of therapy there is a likelihood of pre-existing drug-resistant mutants in the lesions, hence antimicrobial resistance (AMR), a likely explanation why current GBT fails. The foregoing clinicopathological features reflect pharmacokinetic (PK) and pharmacodynamic (PD) factors that affect therapy outcomes; however, PK/PD science was not considered in designing GBT. These PK/PD and clinicopathological features, and human-like intralesional PKs were made part of the basic design of the HFS-MAB, consistent with the FDA proposal of NAMs “with high relevance to human biology” (6). Here, we performed a systematic review of this NAM, for lessons learned, in order to improve testing of new drugs for MAB-LD.

## METHODS

### Objectives and study question**s**

The first objective was to perform a systematic review and quantitative analysis to validate and benchmark the HFS-MAB for drug development, for presentation to the FDA and EMA (6, 7, 22–28). The second objective was to rank new/repurposed drugs based on the extent of microbial kill, to design a new regimen. The third objective was to use the systematic review PK/PD findings to update dosing of new formulations and their susceptibility breakpoints.

### Minimal criteria for accepting studies

Antimicrobial PK/PD science examines the relationship between drug exposure at site of infection (lung lesions) versus PD (microbial kill or AMR) and has been the foremost approach to abrogating AMR emergence. Microbial effect modeling is performed using the 4-parameter inhibitory sigmoid E_max_ model with the following parameters: [1] the E_con_ is the bacterial burden in non–treated controls, [2] E_max_ is maximal effect or efficacy, [3] H is the Hill slope, and [4] EC_50_ is the drug exposure mediating 50% of E_max_ or potency (29). The equation is as follows;

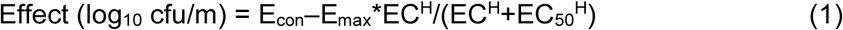

Since this model has 4 parameters, at a minimum 4+1 doses or exposures are required for the model to have a unique solution, and this is the minimum definition for an acceptable exposure-response HFS-MAB study. The antibiotic resistance arrow of time has 3 parameters, so the 5 parameters will be enough as well to identify the drug exposure associated with AMR suppression (30, 31). Finally, by definition, a PK/PD study must have a PK arm-, hence no static concentrations of drug. Therefore, we only accepted studies that measured drug concentrations in the central compartment of the HFS-MAB, to confirm the dynamic concentration-time profiles.

### Quality scores

Recently we developed a quality scoring tool for *M. avium complex* (MAC) HFS and animal PK/PD models, which we used here without modification. The tool is shown and explained in **Supplementary Methods**. The score is categorized as a high score if >20, good score if 15-20, adequate if 10-14, poor quality if 5-9, and deficient and unreliable if <5.

### Literature search

Two authors (TG and SS) searched PubMed, Google Scholar, Interscience Conference on Antimicrobial Agents and Chemotherapy (ICAAC) abstracts, and ID Week abstracts, for studies published through May 1, 2025. The last search was on May 19, 2025. We utilized the medical subject heading (MeSH) “hollow fiber” AND “*Mycobacterium abscessus*” for our search. We also included manuscripts we had submitted to journals (our internal databases), even if not yet on PubMed. The two authors independently extracted the data into the prespecified table format, and then each independently scored the studies for quality criteria, as described below. Consensus was reached for study inclusion after the discussion of each study. There was no exclusion of articles by language. Bias minimization was according to Preferred Reporting Items for Systematic Reviews and Meta-Analyses (PRISMA) [https://www.prisma-statement.org] (32).

### Data and outcomes collected

Data collected included number of HFSMAB units, replicates used, number of doses tested, type of study design (exposure-effect or dose fractionation or factorial design), number of non-American Type Culture Collection (ATCC) clinical isolates tested, and number of doses in MCEs. The outcomes recorded were day zero microbial burden (*B0*) achieved, colony forming units (cfu) per mL, microbial kill below *B_0_*, PK/PD parameter linked to efficacy and AMR suppression, EC_80_, and *in silico* dose-ranging to identify optimal doses in published MCE.

### Data synthesis and analysis

First, we performed a qualitative analysis of lessons learnt, including PK/PD parameters linked to effect, emergence of AMR, and duration of the HFS-MAB experiments. This approach is descriptive. Next, we performed a quantitative analysis that calculated the rate at which a drug killed below *B_0_* when at E_max_ in log_10_ CFU/mL as a fold difference with the three-drug GBT. Since one of the major drivers of quality score was the number of non-ATCC isolates used, and the standard ATCC isolates over-exaggerate efficacy, microbial kill was inversely weighted by the number of non-ATCC isolates used in the study. We also captured the EC_80_ PK/PD target for each study, and PK/PD susceptibility breakpoints from published MCE, where available. The reliability of each study was qualified by the quality score.

### MCE for inhalational therapy

For those studies where the inhibitory sigmoid E_max_ model-based PK/PD target exposures such as the exposure mediating 80% of E_max_ (EC_80_) had not been identified, we calculated those to enrich the literature. These target PK/PD exposures must be achieved at the site of infection to kill the pathogens. A recent approach to achieve the PK/PD exposure target for cefoxitin, tigecycline, amikacin, and imipenem/cilastatin (herein “imipenem”), hitherto administered intravenously, is inhalational therapy formulations (33–41). Therefore, we performed MCE to identify inhalational therapy doses for drugs achieving or exceeding EC_80_ in epithelial lining fluid (ELF). Based on receptor theory and antibiotic as a ligand occupying its target (e.g. clarithromycin binding to subunit 50S of the MAB ribosome), a saturable process, a drug cannot kill more than its E_max_ even if the exposure is increased, therefore, the effect of inhalational doses was not to improve E_max_ but the probability of target attainment (PTA) (42, 43). The population PK model data in the domain of input were selected from the literature and are shown in **Supplementary Methods** (34–41). For imipenem we created an inhalational PK model based on ELF and serum concentrations reported by Badia et al (40). The tigecycline model was published in our recent paper of pulmonary MAC (38). For tigecycline, MICs were published by Terschlüsen e al, but for all other agents MICs were published by Ruedas-Lopez et al (3, 44).

## RESULTS

### Literature search

The literature search is shown in **Figure 1**. Out of 26 publications, 12 were classified as true HFS-MAB PK/PD studies (8, 20, 45–53) including one of our studies out for review, while 14 were not (5, 30, 34, 54–64). The most common venue (4/12) for publication was Antimicrobial Agents and Chemotherapy, which were all 4 first publications (8, 45–47). Ten HFS-MAC studies were monotherapy for amikacin (twice), tigecycline, moxifloxacin, omadacycline, imipenem, apramycin (compared to amikacin), epetraborole, ceftaroline-avibactam, and sulbactam-durlobactam alone and with ceftriaxone (double β-lactam). Two were combination therapy for GBT, and two consisted of both monotherapy and combination therapy studies. Studies were performed in four different laboratories, mostly with the ATCC reference isolate 19977 (ATCC#19977), and only recent studies included clinical isolates.

**Figure 1.**
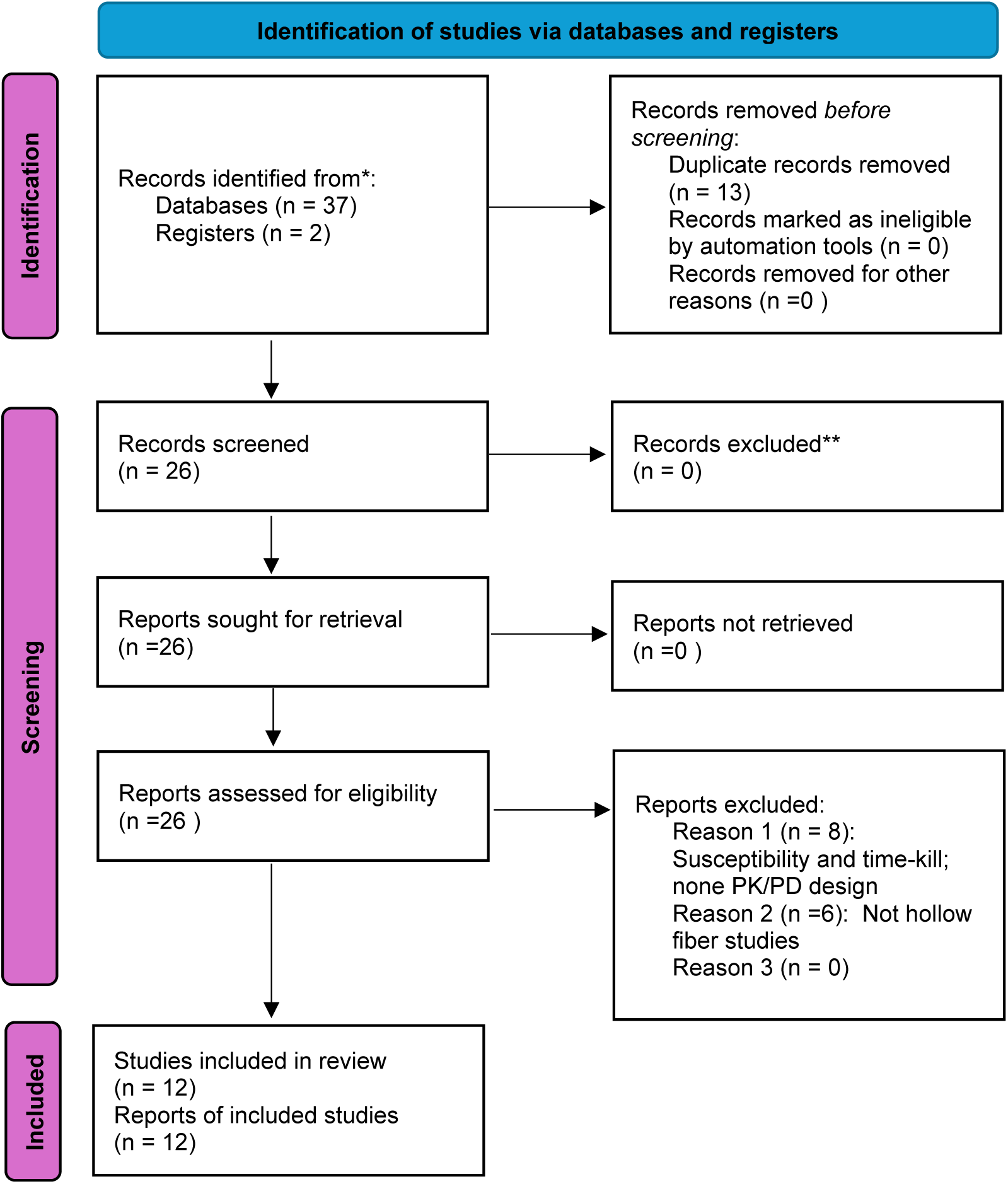
PRISMA Flow Diagram for Studies. The template used was from Page et al (32). The most important reasons for exclusion of studies were duplicate records between databases (n=13), reports of susceptibilities and time-kill studies (n=8), and non-hollow fiber designs.

### Quality scores

Quality scores for the studies are shown in **Figure 2A**. The mean score for monotherapy studies was 13.7±1.07 out of 24 which is in the adequate category, while that for combinations was 9.75±7.41 out of 21, which is in the poor category. The most deficient scoring criteria were for the non-inclusion of non-ATCC isolates, followed by the lack of replicates. In a correlation study to identify drivers of the score (Pearson r>0.5 or <-0.5), the statistically significant scoring criteria were those shown in **Figure 2B**. **Figure 2B** shows that the quality score was driven by [1] failure to utilize a *B_0_* similar to that in patient lesions, a measure of “relevance to human biology” (6), [2] low number of exposures tested, and [3] lack of MCE to identify to identify clinical dose, which is the point of PK/PD studies.

**Figure 2.**
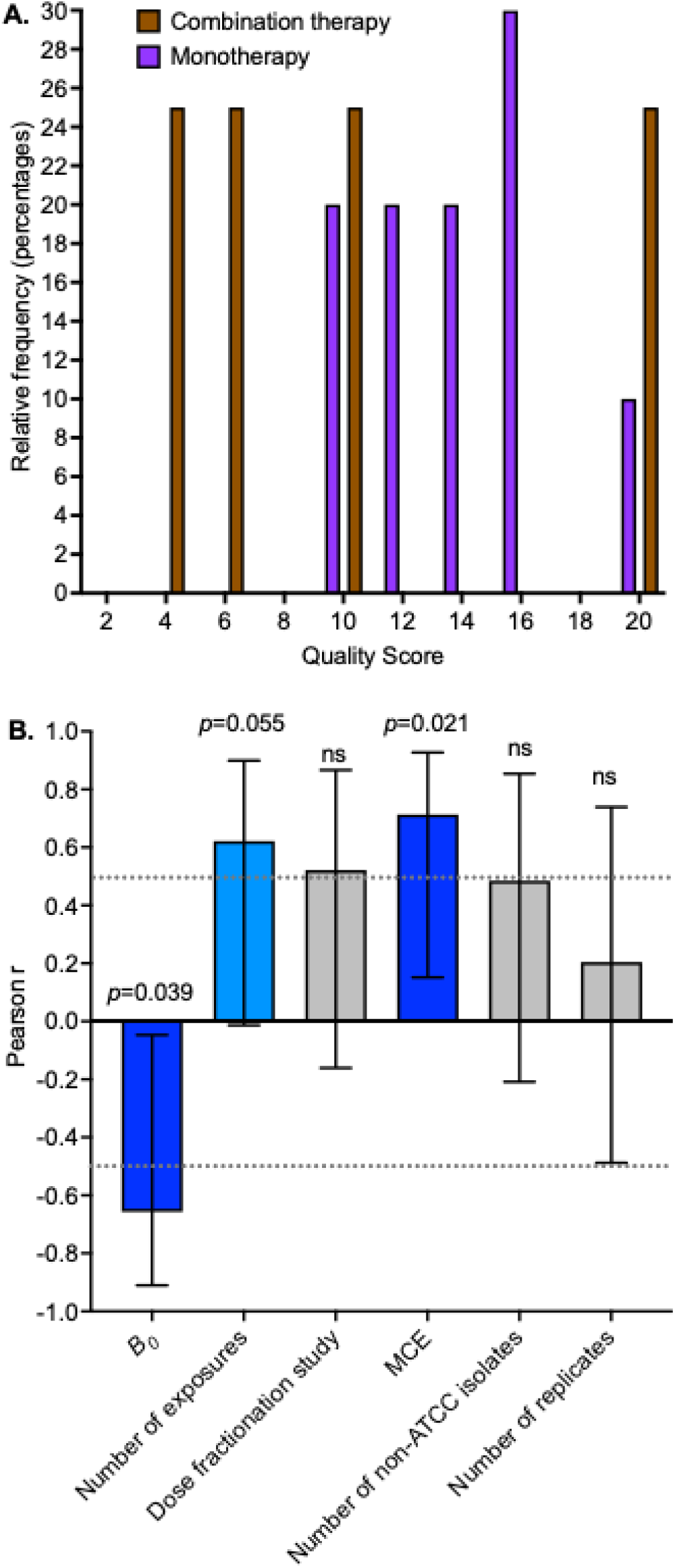
Quality Score of HFS-MAB Studies in the Literature. **A.** Distribution of quality scores shows that the majority of studies had a score below the adequate category (score<15), with at least one study in the deficient category (score <5). **B.** Error bars are 95% confidence intervals. Hatched line is the Spearman r of 0.5 or -0.5, which we used as threshold choosing drivers of quality score. Statistically significant drivers of the scores are shown in shades of blue.

### Qualitative Systemic analysis findings and real-world evidence (RWE)

The studies, qualitative findings, and their quality scores are shown in **Table 1**. The first HFS-MAB was for amikacin and was published in 2015 (8). Since then, there have been two other amikacin studies which demonstrated a 10-fold difference in the extent of microbial kill (8, 49). One study found C_max_/MIC linked effect with an EC_80_ of 3.2, while the other demonstrated %T_MIC_ linked effect with an EC_80_ of 40% (8, 49). There were three GBT studies which we compared to 34% SCC in patients in **Table 1**; two demonstrated virtually the same microbial kill below *B_0_*, while the third study demonstrated no microbial kill below *B_0_* (20, 47, 51). Nevertheless, all three GBT studies demonstrated poor efficacy in HFS-MAB, as encountered in patients. Drug administration was either for 14 days or 21 days. AMR developed in all studies. One study used the term “tolerance” for the AMR; however, the authors did not investigate if this was true tolerance or constitutive induction of efflux pumps which is part of the antibiotic resistance arrow of time (30). Overall, the monotherapy studies demonstrated that the HFS-MAB is tractable as a tool for [1] exposure-effect studies, [2] dose fractionation studies, and [3] comparison of combination regimens, with the PD output of both microbial kill and AMR.

**Table 1.**
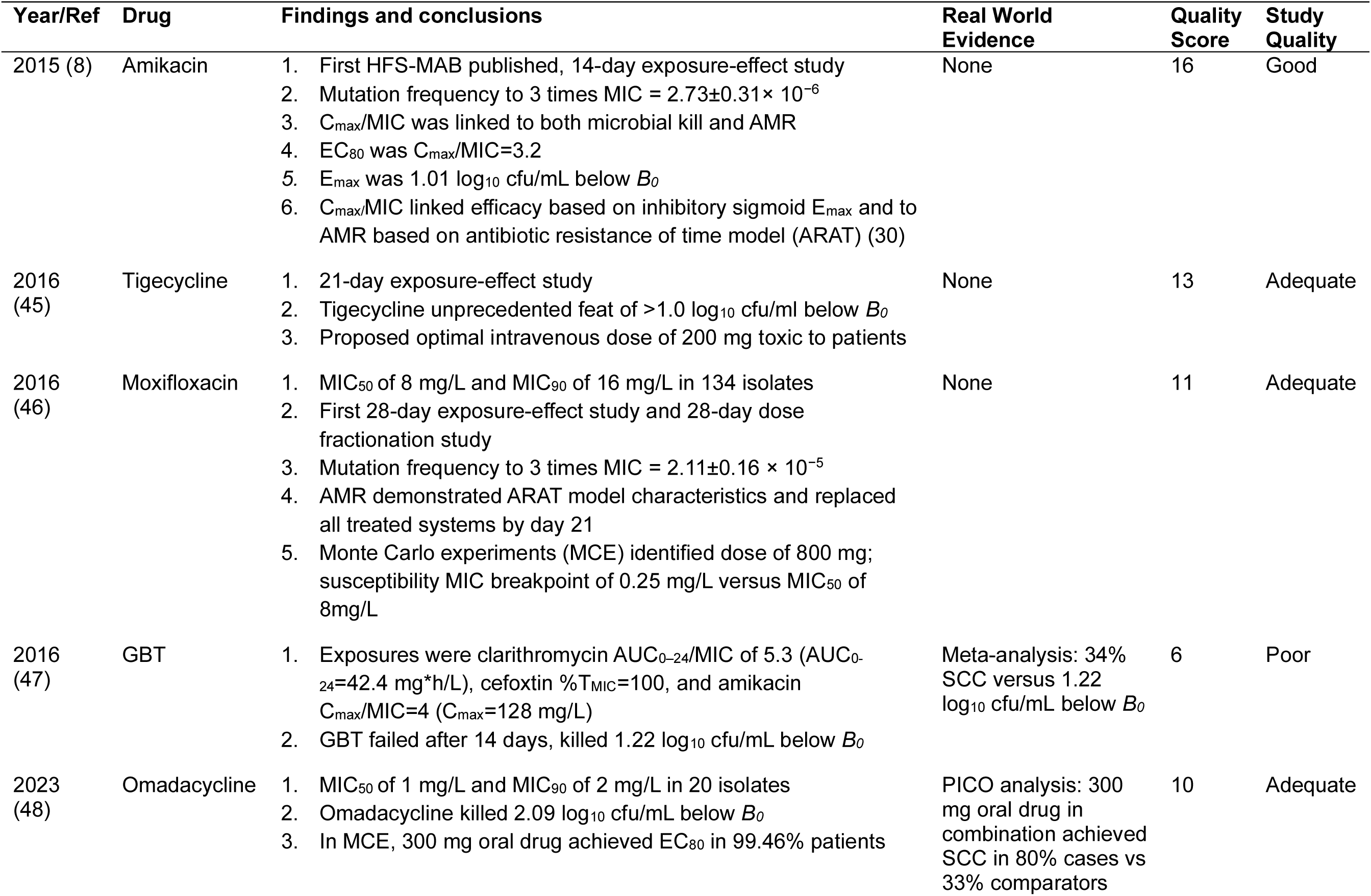
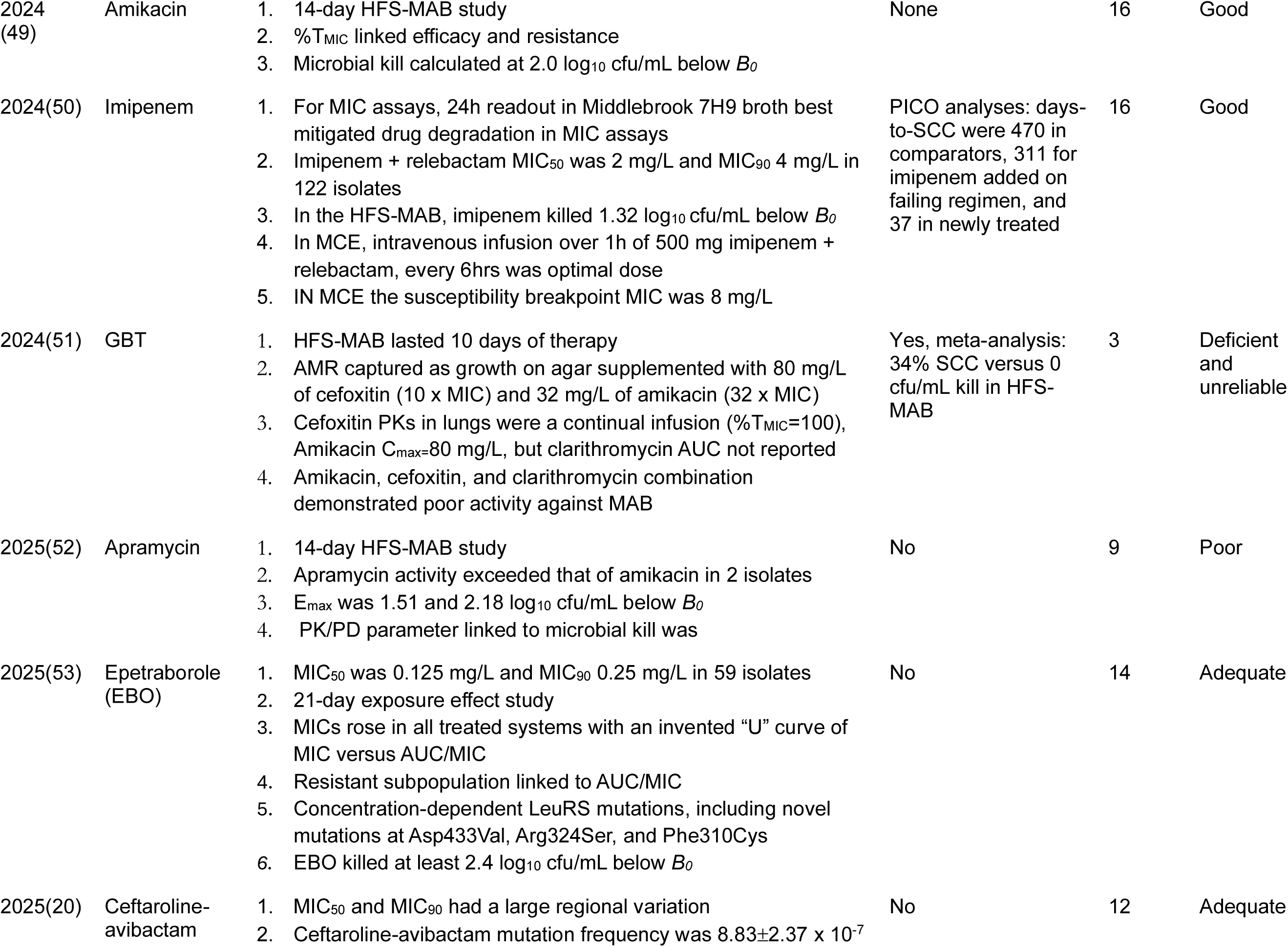
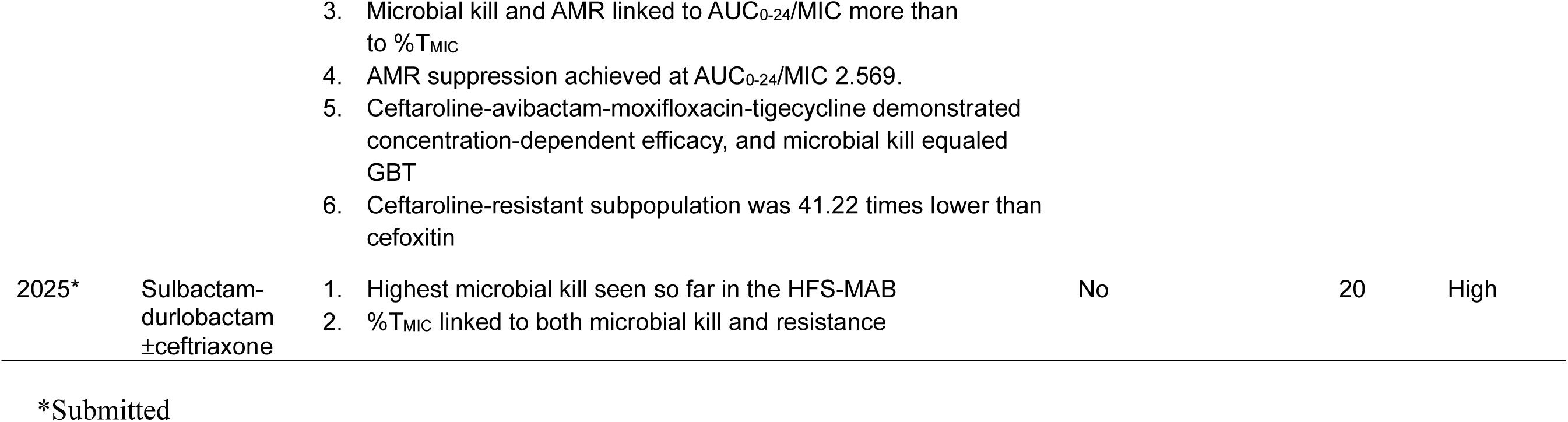
Qualitative Systematic Analyses Findings and Quality of Studies by Year of Publication.

An important contribution in **Table 1** was that of two studies that also presented RWE clinical data versus the HFS-MAB output (48, 50). Guidelines use the PICO (Population, Intervention, Comparators, Outcomes) approach for RWE, an approach that was used in tandem with the two HFS-MAB studies (4, 48, 50). In the first, omadacycline killed 2.08 log_10_ cfu/mL below *B_0_* in the HFS-MAB (that is 7.2-fold better than GBT in the same model) (48). The HFS-MAB-derived EC_80_ was used in MCEs and the omadacycline standard 300 mg/day oral dose was identified as optimal. In the PICO analyses omadacycline-based combination therapy achieved SSCC that was 8-fold better than in comparator patients on GBT that also included tigecycline. In addition, the faster time to SCC was confirmed in prospective trialing in one patient for salvage therapy (48). Similarly, imipenem killed 1.23 log_10_ CFU/mL below *B_0_* in the HFS-MAB, MCEs identified an optimal dose and dosing schedule, and RWE revealed that imipenem-based combination regimens achieved a SCC of 83% in treatment naïve patients, 43% in treatment experienced patients, versus 29% in comparators on GBT (50). The learnings were that biologic activity in the HFS-MAB, and microbial kill below *B_0_* could be translated to patients, based on RWE.

### Quantitative analysis and ranking of drugs by kill below *B_0_*

Given the importance of microbial kill below *B_0_* we ranked all drugs by efficacy (Δ cfu/mL), with GBT as the referent effect, in **Table 2**. Sulbactam-durlobactam-ceftriaxone was ranked highest, epetraborole was ranked second, and omadacycline was ranked third. The PK/PD target exposures for dose selection are shown in **Table 2**.

**Table 2.** Quantitative Systemic Analysis Findings and Ranking of Efficacy of Drugs for Monotherapy.

| Drug (Reference) | $E_{\max}$ ($\log_{10}$ cfu/mL)<br>kill below $B_0$ | Fold of 3-Drug<br>GBT kill below $B_0$ | Rank | PK/PD Target | Dose Schedule<br>(oral and IV) | PK/PD MIC<br>breakpoint (mg/L) |
| --- | --- | --- | --- | --- | --- | --- |
| **MONOTHERAPY** |  |  |  |  |  |  |
| Sulbactam–<br>durlobactam/ceftriaxone | 3.47 | 176.8 | 1 | %T<sub>MIC</sub>=47% | Twice daily |  |
| Epetraborole (53) | 2.44 | 14.76 | 2 | AUC<sub>0-24</sub>/MIC=202.73 | Once daily or twice daily | 0.5 |
| Omadacycline (48) | 2.08 | 7.24 | 3 | AUC<sub>0-24</sub>/MIC=23.76 | Once daily | 1.0 |
| Apramycin (52) | 2.07 | 6.85 | 4 | Too few exposures to<br>calculate PK/PD target | Not clear | None |
| Amikacin (average of 4<br>weighted by score) (8, 49, 52) | 1.6 | 2.42 | 5 | C<sub>max</sub>/MIC=3.2;<br>%T<sub>MIC</sub>=40% | Liposomal amikacin,<br>once daily to once<br>weekly | 8 |
| Tigecycline (45) | 1.3 | 1.47 | 6 | AUC<sub>0-24</sub>/MIC=36.65 | Once daily | 0.5 |
| Imipenem/relebactam (50) | 1.23 | 1.00 | 7 | %T<sub>MIC</sub>=48% | 1h infusion every 6hrs | 8 |
| Ceftaroline/avibactam (20) | 0.69 | 0.29 | 8 | AUC<sub>0-24</sub>/MIC= 9.84 | Twice daily |  |
| Moxifloxacin (46) | 0 | 0 | 9 | AUC<sub>0-24</sub>/MIC=102.11 | Once daily | 0.25 |
| **COMBINATION THERAPY** |  |  |  |  |  |  |
| GBT 1 (47) | 1.23 | 1 | Ref | Not applicable | Different for each drug | Not applicable |
| GBT 2 (51) | 0 | 0 |  | Not applicable | Different for each drug | Not applicable |
| GBT 3 (20) | 1.20 | 0.94 |  | Not applicable | Different for each drug | Not applicable |
| Ceftaroline/avibactam-<br>moxifloxacin-tigecycline (20) | 0.48 | 0.18 |  | Not applicable | Different for each drug | Not applicable |

### MCE with new inhalational formulations

Next, given the availability of the inhaled formulations for tigecycline, amikacin liposome inhalation suspension (ALIS), imipenem, and cefoxitin, we performed additional MCE for these formulations. The population PK parameter estimates in the domain of input derived from the literature are compared to those in 10,000 MCE subjects in **Table 3** (34–41).

**Table 3.** Population Pharmacokinetic Parameter Estimates.

|  | Subroutine PRIOR<br>Estimate | %CV | 10,000 virtual subjects<br>Estimate | %CV |
| --- | --- | --- | --- | --- |
| **Amikacin liposome inhalation suspension (ALIS)** |  |  |  |  |
| Clearance in ELF compartment in L/h | 0.16 x 10<sup>-2</sup> | 71.8 | 0.16 x 10<sup>-2</sup> | 70.82 |
| Volume in ELF compartment in L | 0.08 | 35.1 | 0.08 | 34.88 |
| Clearance in central compartment in L/h | 0.02 | 71.8 | 0.01 | 72.29 |
| Volume in central compartment in L | 0.05 | 65.09 | 0.05 | 65.1 |
| Clearance in peripheral compartment in L/h | 1.99 | 30.24 | 1.05 | 29.78% |
| Volume in peripheral compartment in L | 3.25 x 10<sup>-4</sup> | 40 | 3.26 x 10<sup>-4</sup> | 39.56 |
| **Cefoxitin** |  |  |  |  |
| Clearance in ELF compartment in L/h | 1.82 x 10<sup>-3</sup> | 55.96 | 1.82 x 10<sup>-3</sup> | 55.96 |
| Volume in ELF compartment in L | 0.02 | 44.87 | 0.02 | 44.41 |
| **Imipenem** |  |  |  |  |
| Clearance in ELF compartment in L/h | 1.54 x10<sup>-1</sup> | 100 | 1.54 x10<sup>-1</sup> | 98.15 |
| Volume in ELF compartment in L | 0.06 | 69.4 | 0.06 | 70.87 |
| Intercompartmental clearance L/h | 7.80 | 110 | 7.82 | 107.6 |
| Peripheral Volume (L) | 11.1 | 56 | 11.03 | 56.49 |

**Supplementary Figure S1** shows the amikacin concentration-time profiles for the 5 ALIS doses and dosing schedules during the first 10 days with last dose on day 7. During the first 72h, the MCE-predicted versus to the median concentrations observed in sputum by Rubino et al overlapped, validating our model (37). The PTAs for all ALIS doses and dosing schedules tested for the C_max_/MIC target of 3.2 was 1.0, up to an MIC of 128 mg/L. **Figure 3A** shows the amikacin PTAs for the 40% T_MIC_ target, and demonstrates that all doses, including 590 mg once a week, achieved or exceeded %T_MIC_ of >40% in more than 90% of patients until an MIC of 128mg/L, except in the lowest dose of 285 mg once a week. Thus, the amikacin PK/PD MIC breakpoint for inhaled ALIS was determined as 256 mg/L, which is higher than for IV dosing in **Table 2**.

**Figure 3.**
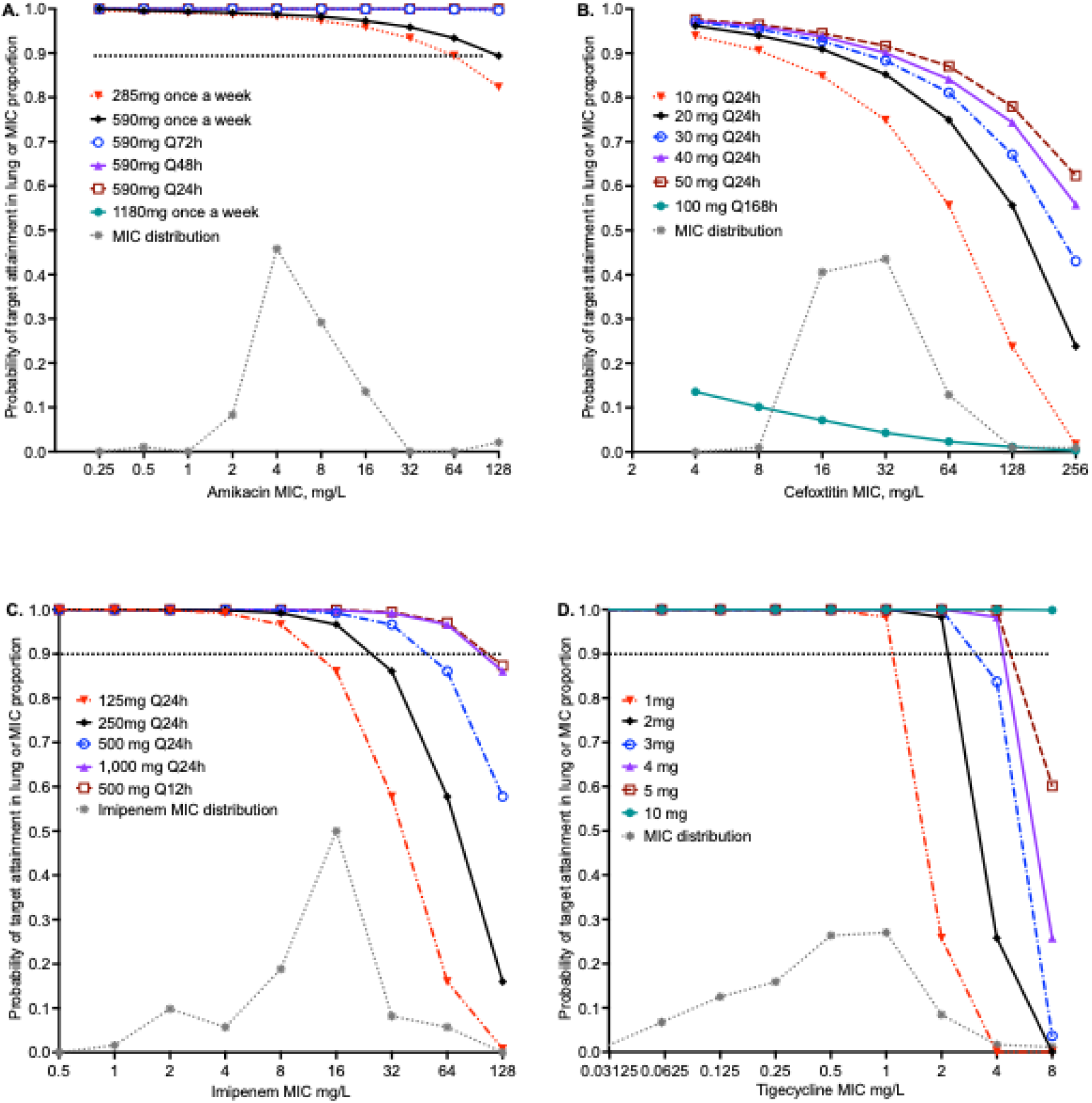
Probability of target attainment for inhaled amikacin, cefoxitin, imipenem, and tigecycline doses and schedules. Probability of target attainment (PTA) for each dose and dosing schedule is shown at each MIC. The symbols indicate PTA estimates for 10,000 patients. The high concentrations achieved by all the drugs achieved in the lungs reflect the low epithelial lining fluid (ELF) volume, compared to the systemic circulation. **A**. Amikacin PTA shows that 590 mg of amikacin liposome inhalation suspension (ALIS) daily, every other day, and every 72h achieved the PTA of ∼100% across the range of MICs measured. The once-a-week regimen falls below 90% at an MIC of 256 mg/L. **B.** Cefoxitin, which has a half-life of about 1h in serum, achieves high concentrations in ELF, and is cleared 6 times slower.(41) We assumed a target of 100% T_MIC_. **C.** The imipenem MIC_90_ was 32 mg/L, and once a day doses ≥500 mg achieved PTA >90% at this MIC. **D.** Low doses of inhaled tigecycline such as 2mg achieved PTA>90% above the MIC_90_.

**Supplementary Figure S2** shows the steady state cefoxitin concentration-time profiles for six inhalation doses. According to the FDA package insert, 1,000 mg of intravenous cefoxitin achieves a C_max_ of 110 mg/L, which means C_max_ with 10 mg would be around 1.1 mg/L versus 516.6 mg/L with the same dose via inhalation route (65). **Figure 3B** shows the PTAs for the cefoxitin doses and schedules, for 100% %T_MIC_ target. The PTA for 50 mg dose was 87% at the MIC_90_ of 64 mg/L, hence, this is the cefoxitin susceptibility MIC breakpoint with nebulization.

**Supplementary Figure S3** shows the steady state imipenem concentration-time profiles for the five inhalation doses and schedules. **Figure 3C** shows the PTAs for 6 doses and schedules, for %T_MIC_ of 48%. The PTA for 250 mg once a day dose was 86% at the MIC_90_ was 32 mg/L, determined as the imipenem susceptibility MIC breakpoint with inhalation.

Tigecycline concentration-time curves were shown in the original paper where model was developed for pulmonary MAC lung disease and are not shown here (38). **Figure 3D** shows the PTAs and demonstrates that even at as low doses as 2 mg, >90% of virtual patients with isolates at the MIC_90_ of 2 mg/L achieved the PK/PD target. At the dose of 4 mg, the tigecycline susceptibility MIC breakpoint after inhalation was 8 mg/L, much higher than the 0.5 mg/L after IV dosing in **Table 2**.

The cumulative fraction of response for the 4 drugs are shown in **Figure 4**. The following inhalational doses were identified as optimal for MAB-LD, 590 mg once weekly for ALIS (**Figure 4A**), 50 mg/day for cefoxitin (**Figure 4B**), 250 mg/day for imipenem (**Figure 4C**), and 4 mg/day for tigecycline (**Figure 4D**).

**Figure 4.**
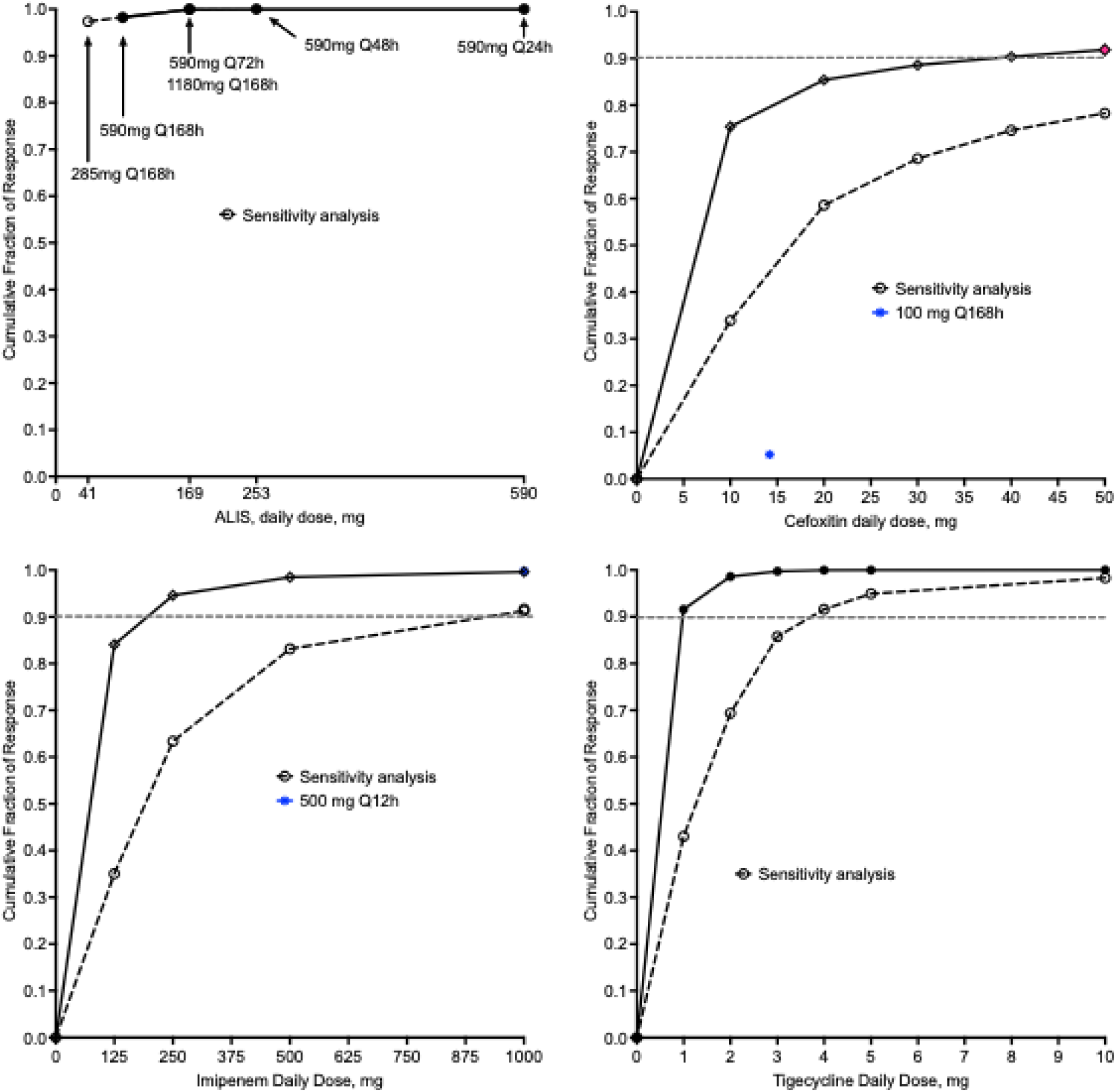
Cumulative Fraction of Response for 10,000 patients at each dose. Sensitivity analysis was made by shifting MICs by two tube dilutions up (that is making the isolates more resistant than observed). Symbols are point estimates for 10,000 virtual subjects. **A.** All doses of amikacin liposomal Inhalation suspension (ALIS) tested achieved a cumulative fraction of response (CFR) of >90%, including the once-a-week dosing. Sensitivity analysis did not change the final estimates. **B.** The cefoxitin dose of 50mg achieved CFR in >90% of patients; sensitivity analysis revealed a CFR of 80%. **C.** The imipenem daily dose 250mg achieved a CFR of >90% but falls to 60% on sensitivity analysis. However, given that unlikely our sensitivity analysis the MICs are likely lower than observed because of imipenem degradation (50), the dose of 250 mg is recommended. **D.** Sensitivity analysis leads to a tigecycline dose of 4mg being sufficient minimal inhaled dose for treatment of pulmonary MAB.

## DISCUSSIONS AND RECOMMENDATIONS

### The HFS-MAB place in the FDA Roadmap and for the EMA

The HFS-MAB fits the FDA roadmap and EMA process to replacing animal testing in preclinical studies with scientifically validated NAMs (6, 7). The current systematic analysis is a step towards that validation. We followed the approach used for qualification of the HFS model for tuberculosis by the EMA and endorsement by the FDA (22–28).

### HFS-MAB model development and quality of evidence-based recommendations

The HFS-MAB quality scores were in the adequate category, which means that they can be improved upon. The two most deficient scoring criteria were the non-inclusion of non-ATCC isolates and lack of replicates. Guidance by the EMA already recommends testing at least 4-5 isolates for more robust PK/PD target setting, which we also recommend for HFS-MAB, and indeed even in animal studies (66). This is also important for the generalization of PK/PD study findings. Here, we propose use of at least two replicates in exposure-effect and dose-fractionation studies, and at least three in all combination therapies.

The quality score was driven by three further factors. The first, failure to utilize a *B_0_* like that in the patient lesions is the easiest to remedy and would not impose any extra costs. Making the *B_0_*“human biology-like” is important because this is one of the most important predictors of efficacy and AMR (10–12, 14–21). The second important score driver was the number of exposures used. Part of our definition of studies to include in this analysis was a minimum of 5 exposures tested. Here the score driver means that it will be advantageous to have more. In practice, the best number of doses is usually seven to eight, including at least one on the steep portion of the dose-response curve. The third score driver was the lack of MCE to identify to identify clinical dose. Why do exposure-response PK/PD studies if the aim is not to find a PK/PD target exposure for use in MCE dose finding? Antimicrobial PK/PD studies are important because they identify the exposure target which determines the optimal dosing regimens for maximal efficacy and minimal AMR. MCEs enable determination of those optimal clinical doses.

### HFS-MAB model tractability

Work in four laboratories shows that this tool can be used by multiple groups. There is a need to further standardize the model as described above. Nevertheless, as the model continues to be improved, here we showed that the HFS-MAB is tractable, with successful completion of exposure-effect and dose-fractionation studies. This model is important for setting PK/PD targets of the new/repurposed drugs, from which clinical doses and susceptibility breakpoints need to be set. Given the variation of kill rates between laboratories, we would like to recommend performance of the 3-drug GBT as a positive control in exposure effect studies. If the GBT and non-treated HFS-MAB behave differently from historical controls, then the study quality would be considered compromised.

The HFS-MAB can also be used for combination therapy studies. This is best done via the factorial design of EC_20_, EC_50_, and EC_80_ exposures, with HFS-MAB replicates. Here, we also show that the biological signal of kill below B_0_ in the HFS-MAB and the MCE predicted dose, correlate well with SCC in patients based on RWE (48, 50). Of course, more work will be required, but this type of translation from HFS-MAB to patients is promising (48, 50). We propose that these currently available data should be presented to regulatory authorities for the context of use discussed here to improve the treatment of MAB-LD, especially considering the FDA modernization act.

### AMR

AMR abrogates effect of GBT in patients with MAC-LD. In the HFS-MAB studies in **Table 1**, AMR also universally terminated microbial kill. MAB has a diverse armamentarium of AMR mechanisms targeting both old/repurposed and new drugs (30, 67). These mechanisms include constitutive induction of efflux pumps in the “antibiotic resistance arrow of time” model, reduced uptake, detoxification, compensatory metabolism shifts, concentration-dependent mutations in antibiotic target sites, and especially multiple drug-modifying enzymes (30, 53, 67). Because repetitive sampling is the *modus operandi* in the HFS-MAB, this allows the tracing of evolution to development of AMR under different drug exposures, over time, as is the case with patients’ sputa, but not possible in animal models. Importantly, the HFS-MAB gives a preclinical drug development tool in which combination therapy can now be tested for the ability to abrogate AMR for MAB-LD, to develop regimens that will not fail.

### HFS-MAB as a ranking tool that informs combination therapy

MAB-LD is an orphan disease. The Orphan Drug Act defines a rare disease as a condition that affects less than 200,000 people, therefore, the number of participants available for MAB clinical trials will be limited. In this context, the HFS-MAB is an agnostic platform to rank monotherapy drugs at optimal exposure for each drug by how well they will perform relative to GBT, at clinical doses identified by MCEs. The three top-ranked drugs (excluding runner-up in the same pharmacophore) can then be combined in a factorial design (using EC_20_, EC_50_, and EC_80_) in the HFS-MAB and compared to GBT. Here, based on the information available to date, these would be sulbactam-durlobactam/ceftriaxone, epetraborole, and omadacycline. The result could be a novel regimen to compare in an open-labeled clinical trial with microbiologic outcome (SCC), with patient-reported outcomes as secondary. Failures of SCC after 6 months of therapy with novel regimens can then be treated with GBT. This avoids the usual tendency to add one new drug to GBT at a time and then switch out components which exposes patients to potentially more toxicity and stretching out trials for many years. This also means that championing of personal favorite regimens for an orphan disease could be replaced by this agnostic tool which ranks drugs based on improvement to GBT, which is what patients want (68).

### HFS-MAB as a tool to identifying experimental doses for the clinic

We examined the doses of 4 drugs that are currently administered intravenously, some administered multiple times a day, and even if once-a-day are associated with considerable toxicity. The HFS-MAB output was used to identify doses for inhalational formulations, and the optimal doses thereof. The ALIS results demonstrated that 590mg a day, the dose used in clinical trials such as the CONVERT study, would achieve the target exposures in >99% of patients (69). Indeed, a once-a-week dosing schedule for the same dose was shown to be optimal as well, which may improve patient compliance. Based on the HFS-MAB and MCEs, we report new doses for cefoxitin, imipenem, and tigecycline inhalational formulations, for testing in the clinic. In summary, HFS-MAB could be useful to identify dosing for new drugs and new formulations prior to phase 3 trials.

### Limitations

First, we introduced a quality score tool which demonstrated that some studies were of poor quality. The poor quality of the pre-clinical work identified in our systemic analysis is more likely a reflection of poor funding options. HFS-MAB studies are expensive, and it costs more to include replicates, to perform standalone dose-fractionation studies, and to generalize findings using multiple clinical MAB isolates. On the other hand, these steps are important for robust PK/PD target exposure identification. Second, the finding that GBT and amikacin-monotherapy studies resulted in different rates of microbial kill based on the laboratory that performed the study, is an important limitation. However, this can be remedied by inclusion of replicates, and use of multiple MAB isolates. Third, in patients 1-2% of bacteria in cavities were shown to be biofilm embedded in the extracellular cavity matrix (18, 70). While biofilm develops in the HFS-MAB, it starts forming around days 3-7, so that the first few days of HFS-MAB consist of mainly bacteria in log-phase growth without biofilm. In the future, PK/PD studies should be started after day 7 to study the biofilm component of MAB-LD.

## Supporting information

Supplementary Methods and Results

## REGISTRATION OF REVIEW

Not registered.

## ACKNOWLEDGEMENT

None.

## CONFLICT OF INTEREST

TG Founded Praedicare Inc, which uses the hollow fiber methodologies for drug development

## FUNDING SOURCE

None.

## DATA AVAILABILITY STATEMENT

The data for the results presented in the manuscript are publicly available.

## ETHICAL APPROVAL

Not applicable.

## AUTHOR CONTRIBUTIONS

Conceptualization and design, TG; Data analysis, TG and SS. TG wrote the first draft of the manuscript. Both authors reviewed and approved the final version of the manuscript.

## Notes

### Funding Statement

This study did not receive any funding.

