## Supplementary Methods and Results for "Systematic and quantitative analyses of hollow fiber model of *Mycobacterium abscessus* lung disease studies"

#### **SUPPLEMENTRAY DATA**

Supplementary Methods

Supplementary References

Supplementary Results Figures

Supplementary Results Tables

#### SUPPLEMENTARY METHODS

#### STUDY QUALITY SCORING

| Criterion | No | 1 | 2 | 3 | 4 | 5 | Total Possible Score |
|---|---|---|---|---|---|---|---|
| **EXPOSURE EFFECT** | | | | | | | |
| *$B_0$ :*<br>0 score if not reported<br>1 if density is reported but is outside the range seen in patients<br>2 if reported and within range encountered in patients<br>3 if both density and total bacterial burden are reported | 0 | 1 | 2 | 3 | | | 3 |
| Number of exposures:<br><5 exposures =0,<br>5 exposures =1<br>6 exposures =2<br>7 exposures =3<br>8 exposures =4<br>At least one exposure of steep portion = 1 extra point | 0 | 1 | 2 | 3 | 4 | 5 | 5 |
| Dose fractionation study was performed: 0 if non, 2 if fractionation for at least 2 dose, 3 if at least 3 doses fractionated | 0 | | 2 | 3 | | | 3 |
| MCE: score by number of doses tested | 0 | 1 | 2 | 3 | 4 | 5 | 5 |
| Number of non-lab-reference (ATCC) isolates for PK/PD target | 0 | 1 | 2 | 3 | 4 | 5 | 5 |
| Number of replicates:<br>One =0<br>2 replicates=1<br>3 replicates in dose response=2<br>If more than 1 replicate for both reference lab strain and the clinical isolates = 3 | 0 | 1 | 2 | 3 | | | 3 |
| **Total Possible Score** | | | | | | | **24** |
| **COMBINATION** (similar to exposure-effect but replace [1] number of exposures, [2] dose-fractionation, and [3] MCE with- | | | | | | | |
| Score:<br>0 if one dose/exposure of each drug was used,<br>3 if at least 3 non-zero exposures were used for each drug,<br>4 if 2 replicates were used, and 5 if at least 3 replicates were used. | 0 | 1 | 2 | 3 | 4 | 5 | 5 |
| Score:<br>0 if there is no synergy or antagonism interaction index<br>3 if there is an interaction index calculated,<br>4 if the optimal exposures of each component for microbial kill calculated,<br>5 if optimal exposures are calculated for both microbial kill and resistance suppression | 0 | 1 | 2 | 3 | 4 | 5 | 5 |
| **Total Possible Score** | | | | | | | **21** |

#### MONTE CARLO EXPERIMENTS (MCE)

##### PK models

All modeling and MCEs were performed using ADAPT 5 software (1). Lung epithelial lining fluid [ELF] was specified as the direct deposition compartment of a bolus (that is as its own compartment), based on PK modeling of inhaled drug concentration data from the literature. First, we generated the population PK model from published data as a one-, two-, or three-compartment model, and then chose the best model using Akaike Information Criteria and Bayesian Information Criteria, as well as parsimony. The best models were then used for MCE and entered into subroutine PRIOR of ADAPT 5.

##### Amikacin Liposome Inhalation Suspension (ALIS)

Rubino et al described population PK parameters in 53 patients, 14 from TR02-112 trial and 39 patients from CONVERT trial, treated with ALIS 590 mg once each day (2). They analyzed the serum concentrations and built a 3-compartment model. They also presented sputum concentrations of amikacin from the two trials, which they did not use for the population PK model. We analyzed the published sputum concentrations with the assumption that sputum concentrations mirror ELF concentrations. Model comparisons were as shown in **Supplementary Methods Table 1**.

**Supplementary Methods Table 1. Amikacin Population Pharmacokinetic Model Comparisons**

| Compartments | Akaike Information Criteria | Bayesian Information Criteria | $r^2$ |
|---|---|---|---|
| One | 1812.16 | 1822.58 | 0.770 |
| Two | 1818.16 | 1836.39 | 0.770 |
| Three | 1741.47 | 1764.91 | 0.958 |

Therefore, the 3-compartment model was used for the Monte Carlo experiments (MCE), with inter-individual variances used being from the Rubino et al model (2).

**Cefoxitin Inhalation**

In rats that received nebulized cefoxitin which were compared to those treated via the intravenous route (IV) (3). After inhalation, the cefoxitin ELF half-life was 1.54h while that in plasma was 1.23 h, basically similar. However, after IV the cefoxitin ELF half-life was 0.19 h and in plasma was 0.23 h. Thus, inhaled formulations resulted in half-life at least 6 times longer than after IV administration (3). This means that the elimination rate ($k_{el}$) in ELF after inhalation is 6-fold lower than after IV administration. After nebulization the ELF AUC was 119,289 h*mg/L versus plasma AUC of 104 h*mg/L, a ratio of 1,147; the LEF/plasma peak concentrations were 1,000-fold, virtually the same as AUCs (3). Given that the $k_{el}$s in ELF and plasma are similar, and:

$$k_{el} = CL/V$$

the higher AUCs and peak concentrations reflect differences in volumes. Moreover, volume scales at a power of 1 between species (4-6).

Cefoxitin plasma/serum based population PKs in critically ill patients with sepsis from *Enterobacteriae* in a study by Chabert et al were best described using a one compartment model with a clearance of 10L/h and a volume of 12L, which was virtually the same 123 obese patients undergoing elective bariatric surgery different study by Novy et al who had a mean clearance and standard deviation of 10.9 ± 6.1 L/h and volume of distribution of 23.4 ± 10.5 L (7, 8). Therefore, we set the cefoxitin ELF volume at 0.02L with an intra-individual % of 44.87% and an ELF clearance of 1.82 x $10^{-3}$ L/h with an IIV % of 55.96%. We tested nebulized doses of 100 mg, 200 mg, 400 mg, 800 mg, and 1,000 mg a day, prepared in 0.9% NaCl. The PK/PD target for cefoxitin has not been identified, and we assumed a worst-case scenario of 100% $T_{MIC}$.

**Imipenem/cilastatin**

For imipenem/cilastatin we created in inhalational PK model based on ELF and serum concentrations reported by Badia et al (9). The model selection was based on results shown in **Supplementary Methods Table 2**:

**Supplementary Methods Table 2. Imipenem Population Pharmacokinetic Model Comparisons**

| Compartments | Akaike Information Criteria | Bayesian Information Criteria | $r^2$ |
|---|---|---|---|
| One | 83.67 | 85.24 | 0.146 |
| Two | 22.24 | 20.67 | 0.99 |
| Three | 26.24 | 23.89 | 0.99 |

**Tigecycline**

The inhalational tigecycline model was developed for our work with pulmonary *Mycobacterium avium complex* disease, and is described in full in that publication (10).

**Supplementary Methods References**

#### SUPPLEMENTARY RESULTS FIGURES

##### Figure S1. Amikacin Liposome Inhalation Suspension (ALIS) concentration time profiles in lung

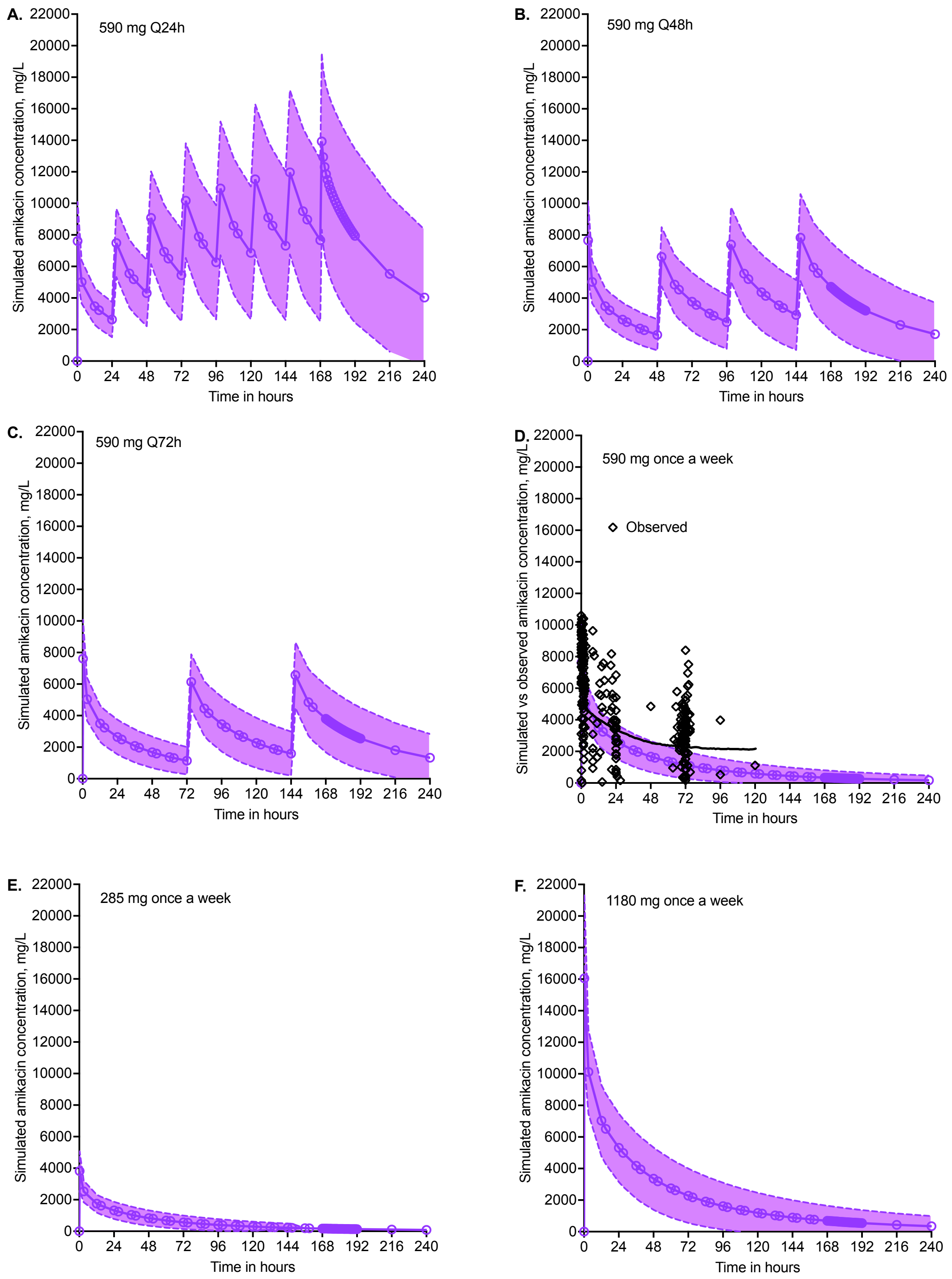

Symbols are mean concentrations in epithelial lining fluid (ELF), while shaded area is standard deviation. **A.** Standard dose administered once each day. **B.** Standard dose administered every other day. **C.** Standard dose administered every 3 days. **D.** Standard dose administered once a week versus concentrations observed in sputum. Our model had slightly faster clearance but overlapped within the first 72h. **E.** Half the standard dose administered once a week. **E.** Two times the standard dose administered once a week.

**Figure S2. Cefoxitin concentration time profiles in lung**

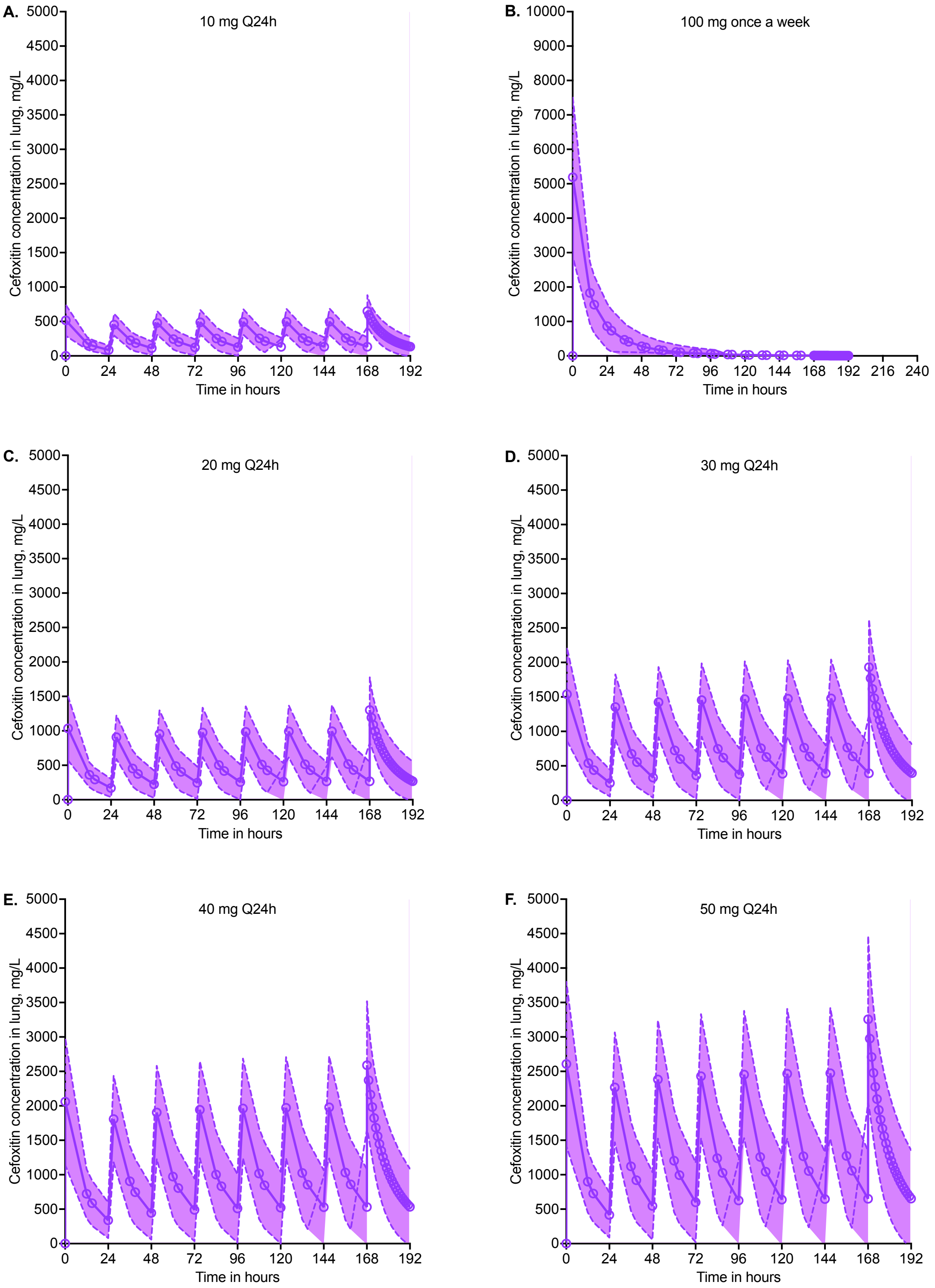


Symbols are mean concentrations in epithelial lining fluid (ELF), while shaded area is standard deviation. All doses were administered once a day.

**Figure S3. Imipenem concentration time profiles in lung**

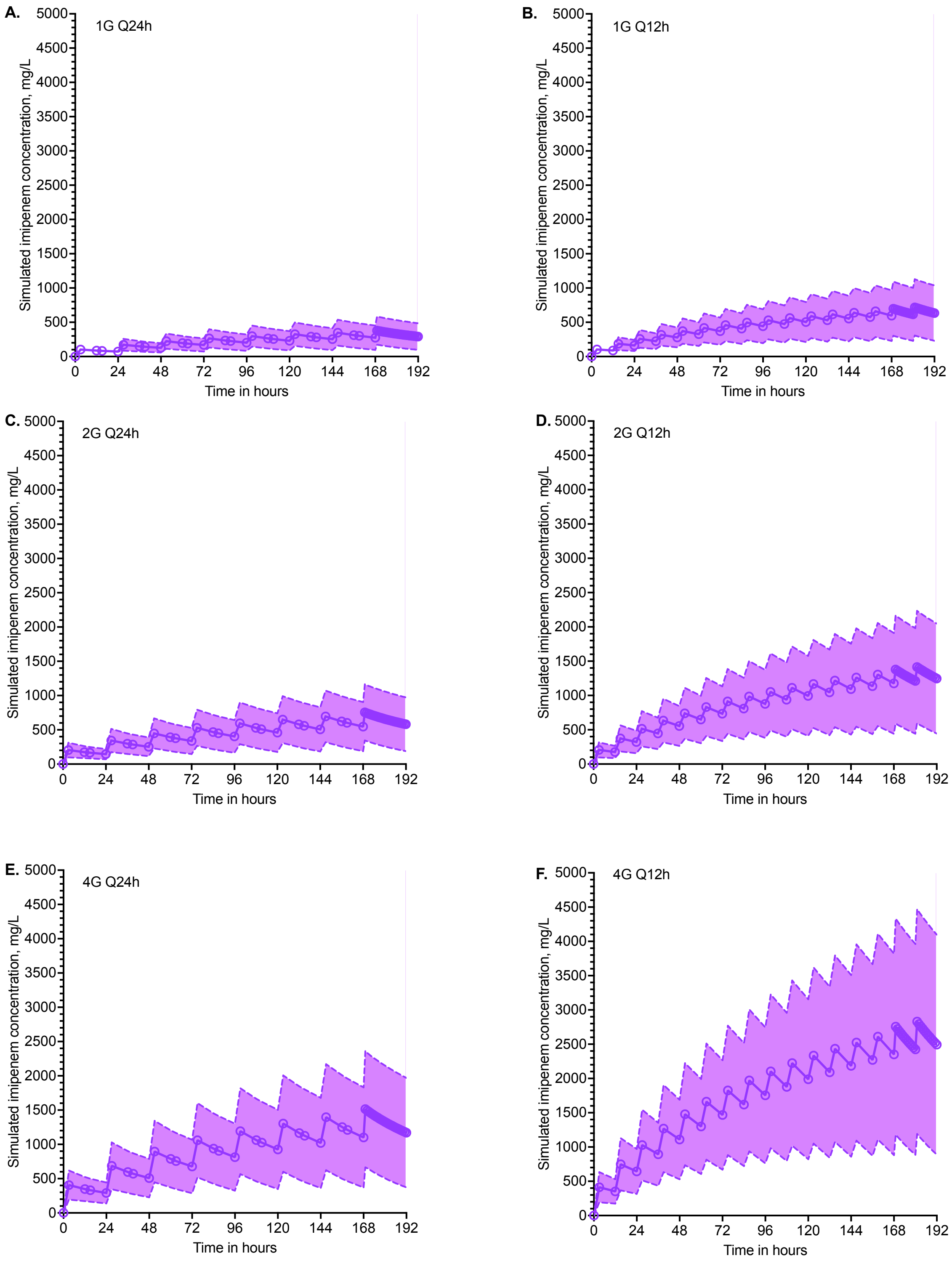


Symbols are mean concentrations in epithelial lining fluid (ELF), while shaded area is standard deviation. **A.** Imipenem 1G once each day. **B.** Imipenem 1G twice a day. **C.** Imipenem 2G once each day. **D.** Imipenem 2G twice a day. **E.** Imipenem 4G once each day. **E.** Imipenem 4G twice a day.

PRISMA 2020 for Abstracts Checklist for Systematic and quantitative analyses of hollow fiber model of *Mycobacterium abscessus* lung disease studies.

| Section and Topic | Item # | Checklist item | Reported (Yes/No) |
|---|---|---|---|
| **TITLE** | | | |
| Title | 1 | Identify the report as a systematic review. | Page 1 |
| **BACKGROUND** | | | |
| Objectives | 2 | Provide an explicit statement of the main objective(s) or question(s) the review addresses. | Lines 28-29 |
| **METHODS** | | | |
| Eligibility criteria | 3 | Specify the inclusion and exclusion criteria for the review. | Lines 29-31 |
| Information sources | 4 | Specify the information sources (e.g. databases, registers) used to identify studies and the date when each was last searched. | In main text |
| Risk of bias | 5 | Specify the methods used to assess risk of bias in the included studies. | Lines 31-32 |
| Synthesis of results | 6 | Specify the methods used to present and synthesise results. | Lines 36-37, 39-40 |
| **RESULTS** | | | |
| Included studies | 7 | Give the total number of included studies and participants and summarise relevant characteristics of studies. | Line 32-34 |
| Synthesis of results | 8 | Present results for main outcomes, preferably indicating the number of included studies and participants for each. If meta-analysis was done, report the summary estimate and confidence/credible interval. If comparing groups, indicate the direction of the effect (i.e. which group is favoured). | Not applicable |
| **DISCUSSION** | | | |
| Limitations of evidence | 9 | Provide a brief summary of the limitations of the evidence included in the review (e.g. study risk of bias, inconsistency and imprecision). | Lines 32-33 |
| Interpretation | 10 | Provide a general interpretation of the results and important implications. | Lines 42-46 |
| **OTHER** | | | |
| Funding | 11 | Specify the primary source of funding for the review. | In main text |
| Registration | 12 | Provide the register name and registration number. | Not registered |

### PRISMA 2020 for Abstracts Checklist for Systematic and quantitative analyses of hollow fiber model of *Mycobacterium abscessus* lung disease studies.

*From:* Page MJ, McKenzie JE, Bossuyt PM, Boutron I, Hoffmann TC, Mulrow CD, et al. The PRISMA 2020 statement: an updated guideline for reporting systematic reviews. BMJ 2021;372:n71. doi: 10.1136/bmj.n71. This work is licensed under CC BY 4.0. To view a copy of this license, visit https://creativecommons.org/licenses/by/4.0/

PRISMA 2020 Checklist for Systematic and quantitative analyses of hollow fiber model of *Mycobacterium abscessus* lung disease studies.

| Section and Topic | Item # | Checklist item | Location where item is reported |
|---|---|---|---|
| **TITLE** | | | |
| Title | 1 | Identify the report as a systematic review. | Page 1 |
| **ABSTRACT** | | | |
| Abstract | 2 | See the PRISMA 2020 for Abstracts checklist. | Yes, attached. |
| **INTRODUCTION** | | | |
| Rationale | 3 | Describe the rationale for the review in the context of existing knowledge. | Lines 82-83 |
| Objectives | 4 | Provide an explicit statement of the objective(s) or question(s) the review addresses. | Lines 86-91 |
| **METHODS** | | | |
| Eligibility criteria | 5 | Specify the inclusion and exclusion criteria for the review and how studies were grouped for the syntheses. | Lines 93-107 |
| Information sources | 6 | Specify all databases, registers, websites, organisations, reference lists and other sources searched or consulted to identify studies. Specify the date when each source was last searched or consulted. | Lines 115-121 |
| Search strategy | 7 | Present the full search strategies for all databases, registers and websites, including any filters and limits used. | Lines 115-121 |
| Selection process | 8 | Specify the methods used to decide whether a study met the inclusion criteria of the review, including how many reviewers screened each record and each report retrieved, whether they worked independently, and if applicable, details of automation tools used in the process. | Lines 115-121 |
| Data collection process | 9 | Specify the methods used to collect data from reports, including how many reviewers collected data from each report, whether they worked independently, any processes for obtaining or confirming data from study investigators, and if applicable, details of automation tools used in the process. | Lines 115-121 |
| Data items | 10a | List and define all outcomes for which data were sought. Specify whether all results that were compatible with each outcome domain in each study were sought (e.g. for all measures, time points, analyses), and if not, the methods used to decide which results to collect. | Lines 127-133 |
| | 10b | List and define all other variables for which data were sought (e.g. participant and intervention characteristics, funding sources). Describe any assumptions made about any missing or unclear information. | Lines 127-133 |
| Study risk of bias assessment | 11 | Specify the methods used to assess risk of bias in the included studies, including details of the tool(s) used, how many reviewers assessed each study and whether they worked independently, and if applicable, details of automation tools used in the process. | Lines 115-121 & Lines 127-133 |
| Effect measures | 12 | Specify for each outcome the effect measure(s) (e.g. risk ratio, mean difference) used in the synthesis or presentation of results. | Lines 135-162 |
| Synthesis methods | 13a | Describe the processes used to decide which studies were eligible for each synthesis (e.g. tabulating the study intervention characteristics and comparing against the planned groups for each synthesis (item #5)). | Lines 135-162 |
| | 13b | Describe any methods required to prepare the data for presentation or synthesis, such as handling of missing summary statistics, or data conversions. | Lines 135-162 |
| | 13c | Describe any methods used to tabulate or visually display results of individual studies and syntheses. | Lines 135-162 |
| | 13d | Describe any methods used to synthesize results and provide a rationale for the choice(s). If meta-analysis was performed, describe the model(s), method(s) to identify the presence and extent of statistical heterogeneity, and software package(s) used. | Lines 135-162 |
| | 13e | Describe any methods used to explore possible causes of heterogeneity among study results (e.g. subgroup analysis, meta-regression). | Lines 135-162 |

PRISMA

### PRISMA 2020 Checklist for Systematic and quantitative analyses of hollow fiber model of *Mycobacterium abscessus* lung disease studies.

| Section and Topic | Item # | Checklist item | Location where item is reported |
|---|---|---|---|
| | 13f | Describe any sensitivity analyses conducted to assess robustness of the synthesized results. | Lines 135-162 |
| Reporting bias assessment | 14 | Describe any methods used to assess risk of bias due to missing results in a synthesis (arising from reporting biases). | Lines 135-162 |
| Certainty assessment | 15 | Describe any methods used to assess certainty (or confidence) in the body of evidence for an outcome. | Introduced a new Quality scores tool (supplementary Methods) |
| **RESULTS** | | | |
| Study selection | 16a | Describe the results of the search and selection process, from the number of records identified in the search to the number of studies included in the review, ideally using a flow diagram. | Figure 1, page 29 |
| | 16b | Cite studies that might appear to meet the inclusion criteria, but which were excluded, and explain why they were excluded. | Line 167-169 |
| Study characteristics | 17 | Cite each included study and present its characteristics. | Table 1 |
| Risk of bias in studies | 18 | Present assessments of risk of bias for each included study. | Table 1 |
| Results of individual studies | 19 | For all outcomes, present, for each study: (a) summary statistics for each group (where appropriate) and (b) an effect estimate and its precision (e.g. confidence/credible interval), ideally using structured tables or plots. | Table 1 |
| Results of syntheses | 20a | For each synthesis, briefly summarise the characteristics and risk of bias among contributing studies. | Table 2 |
| | 20b | Present results of all statistical syntheses conducted. If meta-analysis was done, present for each the summary estimate and its precision (e.g. confidence/credible interval) and measures of statistical heterogeneity. If comparing groups, describe the direction of the effect. | Table 1, Table 2, Table 3, Figure 3 and Figure 4 |
| | 20c | Present results of all investigations of possible causes of heterogeneity among study results. | Table 1, Table 2 |
| | 20d | Present results of all sensitivity analyses conducted to assess the robustness of the synthesized results. | Figure 3 |
| Reporting biases | 21 | Present assessments of risk of bias due to missing results (arising from reporting biases) for each synthesis assessed. | Figure 2 |
| Certainty of evidence | 22 | Present assessments of certainty (or confidence) in the body of evidence for each outcome assessed. | Figure 3 |
| **DISCUSSION** | | | |
| Discussion | 23a | Provide a general interpretation of the results in the context of other evidence. | Lines 269-357 |
| | 23b | Discuss any limitations of the evidence included in the review. | Lines 359-372 |
| | 23c | Discuss any limitations of the review processes used. | Lines 359-372 |
| | 23d | Discuss implications of the results for practice, policy, and future research. | Lines 269-357 |

PRISMA 2020 Checklist for Systematic and quantitative analyses of hollow fiber model of *Mycobacterium abscessus* lung disease studies.

| Section and Topic | Item # | Checklist item | Location where item is reported |
|---|---|---|---|
| **OTHER INFORMATION** | | | |
| Registration and protocol | 24a | Provide registration information for the review, including register name and registration number, or state that the review was not registered. | Not registered |
| | 24b | Indicate where the review protocol can be accessed, or state that a protocol was not prepared. | Not applicable |
| | 24c | Describe and explain any amendments to information provided at registration or in the protocol. | Not applicable |
| Support | 25 | Describe sources of financial or non-financial support for the review, and the role of the funders or sponsors in the review. | Line 378 |
| Competing interests | 26 | Declare any competing interests of review authors. | Line 376-377 |
| Availability of data, code and other materials | 27 | Report which of the following are publicly available and where they can be found: template data collection forms; data extracted from included studies; data used for all analyses; analytic code; any other materials used in the review. | Made available to editors |

*From:* Page MJ, McKenzie JE, Bossuyt PM, Boutron I, Hoffmann TC, Mulrow CD, et al. The PRISMA 2020 statement: an updated guideline for reporting systematic reviews. BMJ 2021;372:n71. doi: 10.1136/bmj.n71.
This work is licensed under CC BY 4.0. To view a copy of this license, visit https://creativecommons.org/licenses/by/4.0/
